# Years of Life Lost Associated with COVID-19 Deaths in the United States During the First Year of the Pandemic

**DOI:** 10.1101/2021.03.02.21252715

**Authors:** Troy Quast, Ross Andel, Sean Gregory, Eric A. Storch

## Abstract

**Background:** Years of Life Lost (YLL) measure the shortfall in life expectancy due to a medical condition and have been used in multiple contexts. Previously it was estimated that there were 1.2 million YLLs associated with COVID-19 deaths in the United States through July 11, 2020. The aim of this study is to update YLL estimates for the first full year of the pandemic.

**Methods:** We employed data regarding COVID-19 deaths in the United States through January 31, 2021 by jurisdiction, gender, and age group. We used actuarial life expectancy tables by gender and age to estimate YLLs.

**Results:** We estimated roughly 3.9 million YLLs due to COVID-19 deaths, which corresponds to roughly 9.2 YLLs per death. We observed a large range across states in YLLs per 10,000 capita, with New York City at 298 and Vermont at 12. Nationally, males had a higher number of YLLs per death than females (9.5 versus 8.8), but there was significant variation in gender differences across states.

**Conclusions:** Our estimates provide further insight into the mortality effects of COVID-19. The observed differences across states and genders demonstrate the need for disaggregated analyses of the pandemic’s effects.

## Introduction

Years of life lost (YLLs) estimates at the population level the number of years that decedents would have lived but for a cause of death. It thus provides insight into the age of those who died and results in higher values when there are a greater number of deaths and/or deaths are more concentrated among the young. The measure has been used to describe all-cause mortality,^1^ as well as for specific causes including non-communicable diseases, ^2-5^ drug misuse, ^6,7^ and suicides.^8,9^ YLLs have been used to characterize deaths due to COVID-19, both in multi-national^10-12^ and single-country^13,14^ analyses.

This short report updates a previous study by the authors of the United States^15^ which estimated 1.2 million YLLs associated with COVID-19 deaths through July 11, 2020. The number of YLLs per death were estimated at roughly 9.3 per death while the top three jurisdictions in terms of total YLLs were New York City, New Jersey, and New York (excluding New York City). This study updates the estimates through the roughly first year of the pandemic, both overall and by jurisdictions and gender.

## Methods

### Data

The data and methods employed follow those used in Quast, et al (2020).^15^ Data regarding COVID-19 deaths was again obtained from the dataset “Provisional COVID-19 Death Counts by Sex, Age, and State” published by the National Center for Health Statistics in the U.S. Centers for Disease Control and Prevention (CDC).^16^ The data reflect the period January 1, 2020 through January 31, 2021 and were obtained on February 3, 2021. We use the term jurisdiction rather than state to refer the geographic entities as the CDC data included observations for the non-state areas New York City and Washington, D.C.. For confidentiality reasons, the number of deaths was suppressed in the dataset for a given jurisdiction/gender/age group if the value ranged from one to nine. Out of 192,545 female deaths, 377 did not include information regarding age. The corresponding values for male deaths was 228,825 and 326. Further, eight deaths had a value of “Unknown” for gender. The suppressed deaths and deaths for which gender was unknown were excluded from the analysis.

Life expectancies by age and gender were obtained from actuarial life tables published by the U.S. Social Security Administration.^17^ The most recent year available was 2017. Appendix Table 1 reports the mapping of age groups used in the COVID-19 deaths data to ages for which the life expectancy was used. The table also reports the approximated life expectancy reported for the specified age. “Annual State Resident Population Estimates” published by the U.S. Census Bureau was the source of our state-level population data.^18^ The data were as of July 1, 2019 and were obtained by gender. Following the COVID-19 deaths data, we estimated the population of New York City and the remainder of New York state separately. We obtained the population of New York City and the percentage of population by gender as of July 1, 2019 from “QuickFacts: New York City, New York” published by the U.S. Census Bureau.^19^

Our sample consisted of 52 jurisdictions: 49 states, New York state excluding New York City, New York City, and Washington, D.C.

### Statistical Analysis

We approximated the age of death for each age group based on the single-age values reported in Appendix Table 1. For each death, we assigned a life expectancy for that gender-age cohort. We obtained population-level YLL estimates by summing the life expectancies for the deaths in the relevant population. Per-capita YLLs were calculated per 10,000 residents for the respective jurisdiction-gender population.

A significant aspect of COVID-19 deaths in calculating YLLs is that many of those who died of the disease had significant pre-existing medical conditions. Such deaths could be considered as displaced mortality in that these individuals on average would likely not reach the full life expectancy reported in actuarial tables. In their forecast model of COVID-19 deaths in the United Kingdom based on data from Italy, Hanlon et al (2020) estimated that the greater pre-existing morbidity of those who died of COVID-19 reduced the estimated YLLs per COVID-19 death from 14 to 13 for men and 12 to 11 for women.^14^ In our analysis we conservatively reduced the expected life expectancy by 25% to reflect the typically greater morbidity of COVID-19 decedents.

## Results

Table 1 reports by jurisdiction the number of deaths and YLLs, both the actual values and those measured per 10,000 capita, ranked in decreasing order by YLLs per 10,000 capita. The roughly 420,000 COVID-19 deaths in the U.S. during the first year of the pandemic translate to nearly 3.9 million YLLS. This corresponds to approximately 120 YLLs per 10,000 capita and 9.2 YLLs per death. New York City has, by far, the highest YLLs per 10,000 capita, at nearly twice the level of the next highest jurisdiction (New Jersey) and nearly 25 times greater than the lowest value (Vermont). California has the highest number of YLLs and represents nearly 11% of the national total, but has only the 31^st^ highest value on a per 10,000 capita basis. Washington, DC has the highest average YLLs per death, indicating a relatively younger age distribution of COVID deaths.

**Table I.** Number of deaths and YLLs by jurisdiction.

| Jurisdiction | Number of Deaths |  | YLLs |  | YLLs per death |
| --- | --- | --- | --- | --- | --- |
|  | Number | Per 10,000 capita | Number | Per 10,000 capita |  |
| Total | 420,682 | 13.0 | 3,861,965 | 119.0 | 9.2 |
| New York City | 23,722 | 28.5 | 248,490 | 298.1 | 10.5 |
| New Jersey | 19,058 | 21.7 | 177,126 | 201.7 | 9.3 |
| Mississippi | 5,592 | 19.0 | 57,627 | 196.0 | 10.3 |
| District of Columbia | 1,046 | 15.0 | 11,746 | 168.7 | 11.2 |
| Arizona | 11,497 | 16.0 | 121,050 | 168.2 | 10.5 |
| Louisiana | 7,428 | 16.2 | 74,444 | 162.2 | 10.0 |
| Alabama | 7,794 | 16.1 | 77,869 | 160.7 | 10.0 |
| New Mexico | 2,960 | 14.3 | 32,630 | 157.3 | 11.0 |
| South Dakota | 1,750 | 20.1 | 13,457 | 154.2 | 7.7 |
| North Dakota | 1,533 | 20.4 | 11,434 | 152.1 | 7.5 |
| New York (excl NYC) | 19,221 | 17.6 | 161,435 | 148.2 | 8.4 |
| Nevada | 4,208 | 13.8 | 44,611 | 146.5 | 10.6 |
| Arkansas | 4,718 | 15.8 | 42,380 | 142.2 | 9.0 |
| Texas | 37,250 | 13.0 | 405,786 | 141.8 | 10.9 |
| Rhode Island | 2,003 | 19.1 | 14,281 | 136.2 | 7.1 |
| Tennessee | 9,391 | 13.9 | 89,341 | 132.4 | 9.5 |
| Illinois | 18,385 | 14.7 | 165,371 | 132.0 | 9.0 |
| Pennsylvania | 21,231 | 16.8 | 163,267 | 128.9 | 7.7 |
| Connecticut | 5,815 | 16.5 | 43,769 | 124.0 | 7.5 |
| Iowa | 5,054 | 16.2 | 37,862 | 121.5 | 7.5 |
| Oklahoma | 5,116 | 13.1 | 47,104 | 120.5 | 9.2 |
| Michigan | 13,070 | 13.2 | 116,195 | 117.6 | 8.9 |
| Massachusetts | 11,258 | 16.5 | 80,040 | 117.3 | 7.1 |
| Indiana | 9,692 | 14.6 | 77,742 | 116.9 | 8.0 |
| Maryland | 7,634 | 12.8 | 69,552 | 116.4 | 9.1 |
| Kansas | 4,041 | 14.0 | 32,696 | 113.6 | 8.1 |
| Georgia | 11,575 | 11.0 | 118,553 | 113.0 | 10.2 |
| Missouri | 8,163 | 13.5 | 67,080 | 110.6 | 8.2 |
| Montana | 1,341 | 12.7 | 11,565 | 109.4 | 8.6 |
| Ohio | 15,923 | 13.8 | 125,780 | 108.9 | 7.9 |
| California | 41,003 | 10.5 | 418,974 | 107.3 | 10.2 |
| Florida | 24,821 | 11.7 | 225,341 | 106.0 | 9.1 |
| Nebraska | 2,432 | 12.7 | 19,909 | 104.3 | 8.2 |
| South Carolina | 5,510 | 10.8 | 51,840 | 101.8 | 9.4 |
| Kentucky | 4,925 | 11.2 | 41,493 | 94.0 | 8.4 |
| Wisconsin | 6,768 | 11.8 | 52,573 | 91.3 | 7.8 |
| West Virginia | 1,835 | 10.3 | 14,892 | 83.9 | 8.1 |
| Delaware | 995 | 10.3 | 8,020 | 83.3 | 8.1 |
| Minnesota | 6,275 | 11.3 | 45,469 | 81.6 | 7.2 |
| Colorado | 5,368 | 9.4 | 45,037 | 79.1 | 8.4 |
| Virginia | 7,200 | 8.5 | 61,059 | 72.4 | 8.5 |
| Idaho | 1,634 | 9.3 | 12,698 | 71.9 | 7.8 |
| Wyoming | 502 | 8.8 | 4,098 | 71.6 | 8.2 |
| Utah | 1,749 | 5.5 | 16,954 | 53.7 | 9.7 |
| New Hampshire | 1,046 | 7.8 | 6,573 | 48.8 | 6.3 |
| Washington | 4,009 | 5.3 | 33,927 | 45.1 | 8.5 |
| North Carolina | 4,073 | 3.9 | 38,584 | 37.2 | 9.5 |
| Oregon | 1,720 | 4.1 | 14,131 | 33.9 | 8.2 |
| Maine | 627 | 4.7 | 4,115 | 30.9 | 6.6 |
| Alaska | 225 | 3.1 | 1,931 | 26.8 | 8.6 |
| Hawaii | 358 | 2.6 | 3,320 | 23.7 | 9.3 |
| Vermont | 138 | 2.2 | 744 | 12.0 | 5.4 |

Figure 1 details YLLs per 10,000 capita by jurisdiction and gender. In every jursidiction, the male value is greater than the female value, which is consistent with YLLs per 10,000 capita for males being roughly a third greater than for females at the national level (136.3 versus 102.3). However, the divergence between the two genders varies considerably by state. In New York City, the male value was nearly 75% greater than the female value, while in Mississippi the male value was only 7% greater.

**Figure 1.**
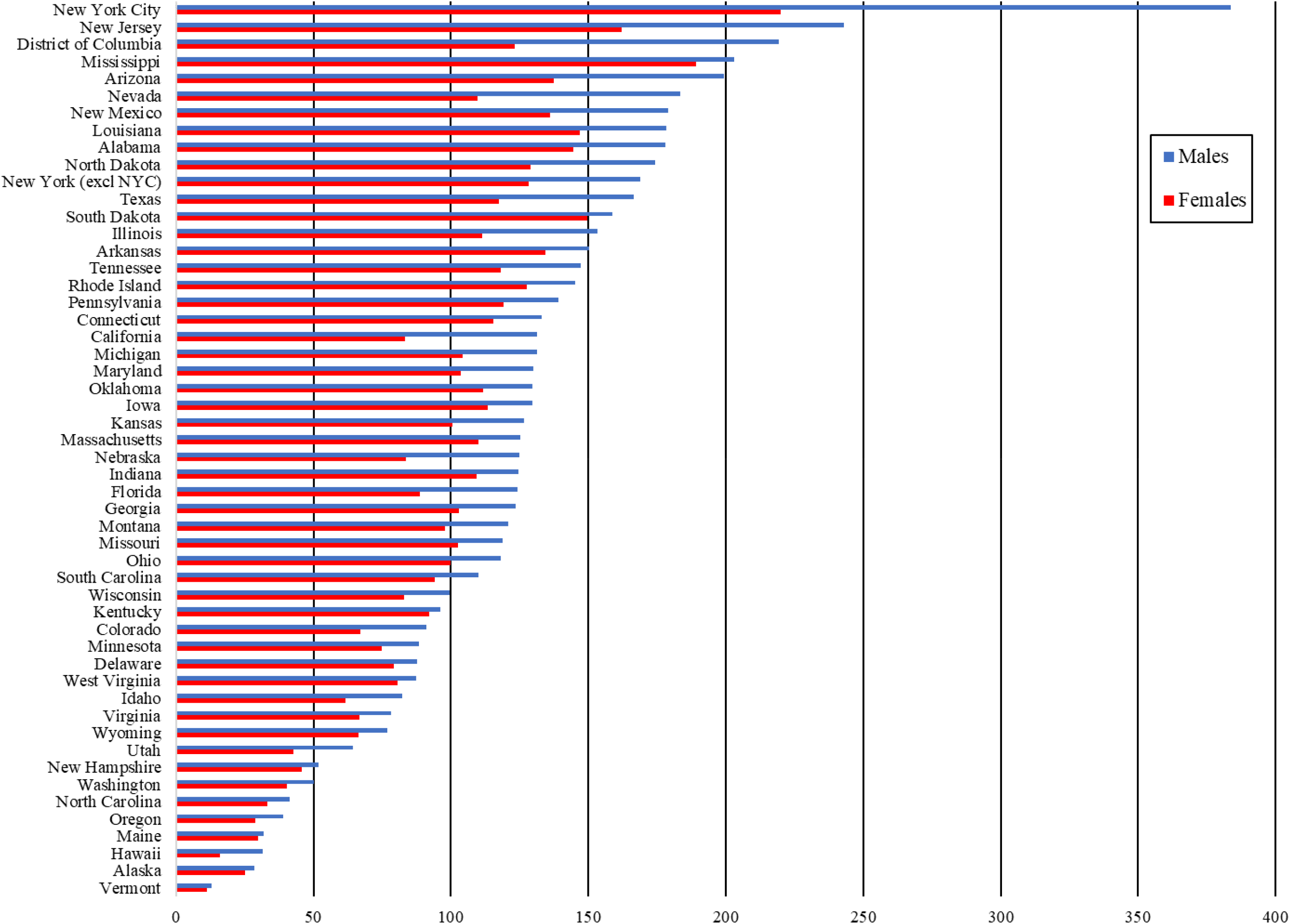
YLLs per 10,000 capita by state and gender.

## Conclusions / Discussion

### The main findings

Over the first year of the pandemic in the U.S., COVID-19 deaths were associated with roughly 3.9 million YLLs. On average, roughly 9.2 years of life were lost per death. We observed dramatic differences across jurisdictions in YLLs per capita. YLLs per capita were greater for females than males, but the extent of the difference varied significantly across jurisdictions.

### What is already known on this topic

Existing studies have found that YLLs associated with COVID-19 deaths in the U.S. are higher than in most other countries. Per-capita U.S. YLLs were found to be roughly 13% greater than in Italy and nearly six times larger than the German value.^10^ An analysis of U.S., U.K., Canada, Norway, and Israel estimated that the U.S. and U.K. YLLs per capita values were the highest among the group and roughly equal.^11^ However, a study of thirty countries with relatively high COVID-19 incidence found that the U.S. had only the eighth highest YLLs per capita.^12^ The only other study specific to the U.S. of which the authors are aware investigated racial and ethnic disparities in YLLs.^13^ The YLLs for blacks and Hispanics were both greater than for whites despite the population of whites being three to four times greater.

### What this study adds

This study updates earlier YLL estimates to include a period of dramatic COVID-19 mortality in the U.S. The number of YLLs estimated in this study are greater by over a factor of three than the number through July 2020. Further, we observed substantial changes in the rankings by jurisdiction relative to our earlier study. For instance, in terms of YLLs per 10,000 capita, Mississippi went from the 14^th^ highest jurisdiction to third.

Our analysis also highlights significant variation in the mortality effects of COVID-19. The dramatic variation in YLLs per 10,000 capita across states highlights differences across geographic regions. We also found substantial heterogeneity in the levels of YLLs per 10,000 capita by gender across states. This finding suggests that differences in the effects of COVID-19 by gender should be investigated at a more local level rather than nationally.

### Limitations

This study shares several limitations with our previous analyis.^15^ We relied on national life expectancy estimates as jurisdiction-level data were not available. Further, our life expectancy estimates are conditional upon an individual reaching the specified age. We had to employ a rough approximation for the pre-existing reduced expected life expectancy of those who died from COVID-19. While our use of a 25% reduction is conservative, especially in light of earlier estimates,^14^ a more precise estimate based on U.S. data would provide greater clarity. Our analysis does not attempt to adjust for quality of life as has been done elsewhere.^11^

The COVID-19 deaths data were provisional and thus incomplete. Estimates based on complete data would result in higher aggregate values, but given our large sample period the differences would likely be relatively minor. There are also concerns as to the accuracy of determinations of deaths caused by COVID-19.^20^ Any undercounting or overcounting of COVID-19 deaths would directly affect our YLLs estimates.

## Conclusions

Our estimates of nearly 3.9 million YLLs and 9.2 YLLs per death in the first year of the pandemic in the U.S. provide another perspective on the striking mortality inflicted by COVID-19. Our analysis also details the substantial variation in mortality effects by state. Further, our gender analysis indicates significant heterogeneity across states. Our findings suggest that attempts to understand the mortality impact of COVID-19 should investigate sub-national regions and sub-populations.

## Data Availability

The CDC death data & Stata programs used are available at https://github.com/troyquast/covid19_ylls_update.

---

**Appendix Table 1.** Mapping of age ranges to single age values and corresponding estimate of life expectancy.

| Age Range | Single Age | Life expectancy<sup>a</sup> |  |
| --- | --- | --- | --- |
|  |  | Female | Male |
| Less than one year | 0 | 80.96 | 75.97 |
| 1 – 4 | 2 | 79.42 | 74.48 |
| 5 – 14 | 10 | 71.50 | 66.57 |
| 15 – 24 | 20 | 61.63 | 56.85 |
| 25 – 34 | 30 | 51.97 | 47.65 |
| 35 – 44 | 40 | 42.47 | 38.56 |
| 45 – 54 | 50 | 33.23 | 29.67 |
| 55 – 64 | 60 | 24.56 | 21.58 |
| 65 – 74 | 70 | 16.54 | 14.39 |
| 75 – 84 | 80 | 9.68 | 8.28 |
| 85+ | 90 | 4.78 | 4.05 |

<sup>a</sup> Estimate prior to any reduction to reflect reduced life expectancy due to greater morbidity of COVID-19 decedents.

## References

1 Vienonen MA, Jousilahti PJ, Mackiewicz K et al. Preventable premature deaths (PYLL) in Northern Dimension partnership countries 2003–13. Eur J Public Health. 2019;29(4):626–630. doi:10.1093/eurpub/cky278

2 Shavelle RM, Paculdo DR, Kush SJ, Mannino DM, Strauss DJ. Life expectancy and years of life lost in chronic obstructive pulmonary disease: findings from the NHANES III Follow-up Study. Int J Chron Obstruct Pulmon Dis. 2009;4:137–148. doi:10.2147/copd.s5237

3 Martinez R, Soliz P, Caixeta R, Ordunez P. Reflection on modern methods: years of life lost due to premature mortality—a versatile and comprehensive measure for monitoring non-communicable disease mortality. Int J Epidemiol. 2019;48(4):1367–1376. doi:10.1093/ije/dyy254

4 Vries E de, Meneses MX, Piñeros M. Years of life lost as a measure of cancer burden in Colombia, 1997-2012. Biomédica. 2016;36(4):547–555. doi:10.7705/biomedica.v36i4.3207

5 Alva ML, Hoerger TJ, Zhang P, Cheng YJ. State-level diabetes-attributable mortality and years of life lost in the United States. Annals of Epidemiology. 2018;28(11):790–795. doi:10.1016/j.annepidem.2018.08.015

6 Imtiaz S, Shield KD, Roerecke M et al. The burden of disease attributable to cannabis use in Canada in 2012. Addiction. 2016;111(4):653–662. doi:10.1111/add.13237

7 Salazar A, Moreno S, Sola HD, Moral-Munoz JA, Dueñas M, Failde I. The evolution of opioid-related mortality and potential years of life lost in Spain from 2008 to 2017: differences between Spain and the United States. Current Medical Research and Opinion. 2020;36(2):285–291. doi:10.1080/03007995.2019.1684251

8 Jung Y-S, Kim K-B, Yoon S-J. Factors Associated with Regional Years of Life Lost (YLLs) due to Suicide in South Korea. IJERPH. 2020;17(14):4961. doi:10.3390/ijerph17144961

9 Izadi N, Mirtorabi SD, Najafi F, Nazparvar B, Nazari Kangavari H, Hashemi Nazari SS. Trend of years of life lost due to suicide in Iran (2006–2015). Int J Public Health. 2018;63(8):993–1000. doi:10.1007/s00038-018-1151-1

10 Mitra AK, Payton M, Kabir N, Whitehead A, Ragland KN, Brown A. Potential Years of Life Lost Due to COVID-19 in the United States, Italy, and Germany: An Old Formula with Newer Ideas. IJERPH. 2020;17(12):4392. doi:10.3390/ijerph17124392

11 Briggs AH, Goldstein DA, Kirwin E, et al. Estimating (quality-adjusted) life-year losses associated with deaths: With application to COVID-19. Health Economics. 2021;30(3):699–707. doi:https://doi.org/10.1002/hec.4208

12 Oh I-H, Ock M, Jang SY, et al. Years of Life Lost Attributable to COVID-19 in High-incidence Countries. Journal of Korean Medical Science. 2020;35(32). doi:10.3346/jkms.2020.35.e300

13 Basset M, Chen J, Krieger N. The unequal toll of COVID-19 mortality by age in the United States: Quantifying racial/ethnic disparities. 2020 Jun 20. https://cdn1.sph.harvard.edu/wp-content/uploads/sites/1266/2020/06/20Bassett-Chen-Krieger_COVID-19_plus_age_working-paper_0612_Vol-19_No-3_with-cover.pdf

14 Hanlon P, Chadwick F, Shah A et al. COVID-19 – exploring the implications of long-term condition type and extent of multimorbidity on years of life lost: a modelling study. Wellcome Open Res. 2020;5:75. doi:10.12688/wellcomeopenres.15849.1

15 Quast T, Andel R, Gregory S, Storch EA. Years of life lost associated with COVID-19 deaths in the United States. J Public Health (Oxf). doi:10.1093/pubmed/fdaa159

16 U.S. Centers for Disease Control and Prevention. Provisional COVID-19 Death Counts by Sex, Age, and State. 2020 Jul 22. https://data.cdc.gov/NCHS/Provisional-COVID-19-Death-Counts-by-Sex-Age-and-S/9bhg-hcku

17 U.S. Social Security Administration. Actuarial Life Table. [undated] <https://w>ww.ssa.gov/oact/STATS/table4c6.html

18 U.S. Census Bureau. State Population by Characteristics: 2010-2019. <https://www.census.gov/data/tables/time->series/demo/popest/2010s-state-detail.html#par_textimage_673542126

19 U.S. Census Bureau. QuickFacts: New York City, New York. <https://www.census.gov/quickfacts/>newyorkcitynewyork

20 Kiang MV, Irizarry RA, Buckee CO, Balsari S. Every Body Counts: Measuring Mortality From the COVID-19 Pandemic. Annals of Internal Medicine. Published online September 11, 2020. doi:10.7326/M20-3100

